# Minimally invasive surgery with a tube-free surgical field for Tetralogy of Fallot repair: A single-center experience

**DOI:** 10.1101/2022.12.05.22283045

**Authors:** Bin Qiao, Zhenglun Alan Wei, Biao Si, Fengquan Zhang, Meng Zhu, Lei Chen, Timothy Slesnick

## Abstract

**Objective:** Several authors have detailed their experiences with small cohorts of patients in light of expanding interest in using minimally invasive surgery (MIS) to treat Tetralogy of Fallot (ToF). The goal of this study was to review an innovative MIS technique that results in a small tube-free surgical field. The technique’s clinical outcomes were examined in the largest cohort to date of patients with ToF treated with an MIS technique.

**Methods:** We reviewed all patients who underwent MIS at a single center between 2013 and 2017. The MIS procedure (including establishment of cavopulmonary bypass) is described. The inter-, peri- and postoperative data are reported and compared with those in the contemporary literature on ToF MIS.

**Results:** A total of 105 patients with ToF were identified. All patients, including 2 under 6 months of age, had good postoperative oxygen saturation (99% [98-100]). The incision size was 3 mm for patients younger than 3 years and 3-5 mm for older patients. No conversions to sternotomy or reinterventions were needed. Postoperative complications occurred in 14 patients (13.3%), including 1 death in the intensive care unit, which was not felt to be cardiac in origin. The primary hospital course metrics were comparable to previously published data.

**Conclusions:** The MIS technique with a tube-free surgical field has been successfully performed in 105 patients. The overall outcomes are favorable, including those for 2 patients younger than 6 months. This innovative MIS could be a promising approach for facilitating ToF repair in patients of all ages.

**Central Picture:** 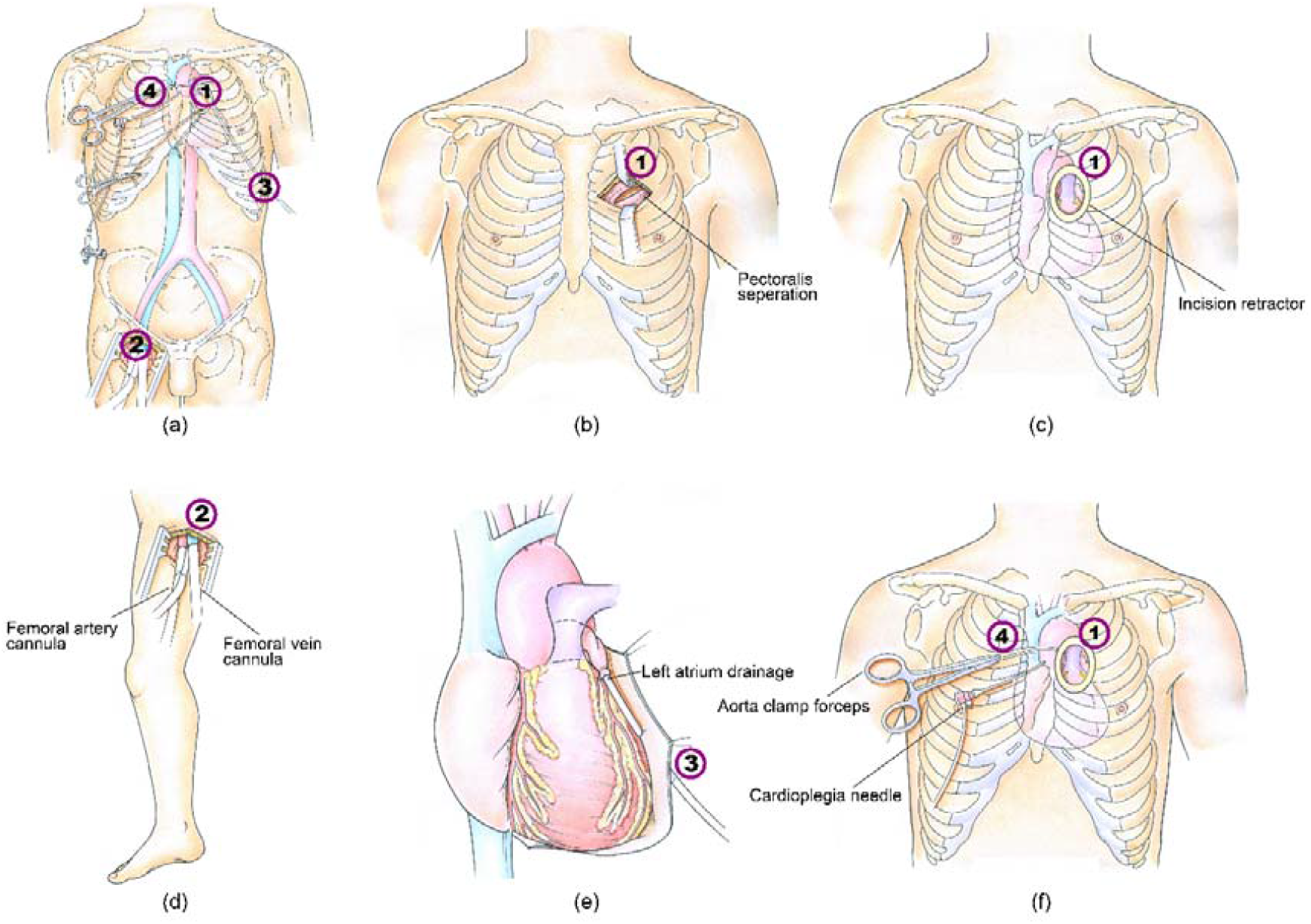

Artist depiction of operative incisions for the proposed minimally invasive surgery.

**Central Message:** This study shows the favorable outcomes of an innovative MIS technique with a tube-free surgical field by reviewing its use in 105 ToF patients, to date the largest cohort undergoing MIS for ToF.

**Perspective Statement:** The proposed MIS technique with a tube-free surgical field presents a promising method for ToF repair; smaller incisions reduce patient pain, facilitate recovery, and enhance cosmesis. This technique achieved overall favorable outcomes for patients with ToF. Also, it can be a good option for early primary ToF repairs.

## BACKGROUND

Tetralogy of Fallot (ToF), the most common form of the cyanotic congenital heart diseases (CHDs), afflicts 1% of newborns each year in the United States.^1^ ToF has 4 primary lesions: right ventricular outflow tract (RVOT) obstruction, right ventricular (RV) hypertrophy, ventricular septal defect, and overriding aorta.^2, 3^ In 1954, the first operation for ToF was successfully conducted using a median sternotomy.^4^ Since then, this surgical technique has been accepted widely as the standard treatment for ToF and generally provides favorable short-term outcomes.^5, 6^

Recently, there has been interest in using minimally invasive surgery (MIS) to treat patients with ToF. MIS may result in (1) smaller incisions, (2) reduced blood loss, (3) shorter in-hospital time, and (4) improved cosmesis,^7, 8^ thereby reducing physical and psychological trauma to the patients.^9^ Though MIS has been used in a range of simple CHDs, including patent ductus arteriosus^10^ and atrial/ventricular septal defect,^11-14^ few studies have described its use for patients with a complex of CHDs such as ToF. Garg et al. treated 11 patients with ToF by using their MIS technique with a transverse sternal split.^15^ Doshi and colleagues reported their experience with a mini-left thoracotomy on 27 pediatric patients.^16^ Lee et al. used a right axillary incision to repair 27 patients with ToF.^17^ Nicholson et al. performed a minimal sternotomy on 27 patients with ToF.^18^ Though each group of authors demonstrated the feasibility of using MIS on patients with ToF, the generalizability of the studies was limited by the small cohort sizes.

Additionally, it is well known that the success of MIS and the associated incision size rely heavily on the cardiopulmonary bypass (CPB) cannulation technique.^19, 20^ Many previous studies used arterial and venous cannulations, as well as aortic clamping, through the main surgery field;^15, 17, 21^ their methods could lead to relatively large incision sizes (3-7 mm). Femoral cannulation has recently become a preferred choice for MIS of many CHDs; however, its application to patients with ToF has thus far been limited to older patients (mean age 13.2 years^16^).

Consequently, there is a pressing need to (1) explore a better cannulation approach that facilitates MIS for patients of all ages with ToF and further reduces surgical trauma and (2) assesses the generalizability of MIS for a relatively large cohort of patients with ToF. To bridge these knowledge gaps, this study details an MIS for ToF with an innovative cannulation approach that results in a small tube-free main surgical field. The clinical outcomes for 105 patients are also reviewed retrospectively, demonstrating the usefulness and efficiency of this MIS in patients with ToF.

## MATERIALS AND METHODS

### Clinical Data Collection

This study retrospectively evaluated 105 patients with ToF from a single institution who underwent MIS for ToF between 2013 and 2017. The preoperative cardiac diagnosis and the size of the pulmonary arteries (PAs) were determined by transthoracic echocardiography for all patients. Further investigation, such as angiography of the femoral artery and vein, was performed as indicated. Associated anomalies were reported. No patient had received a palliative shunt prior to ToF repair. The institutional ethics committee approved the study (RE2012-16). Written informed consent was obtained from the parents or guardians.

### Study Variables

In addition to baseline patient characteristics, recorded intraoperative variables included the duration of the surgical procedure, CPB, cross-clamping, and femoral artery and vein cannulation, as well as anesthetic methods, regional cerebral oxygen saturation, colloid osmotic pressure, and lactic acid levels. The outcome data were chest drainage volume, length of stays in the intensive care unit (ICU) and in the hospital, duration of mechanical ventilation, additional interventions, and drug regimens in the ICU. Other adverse events, such as death and surgical complications, were recorded if they occurred.

Continuous variables were presented in the form of mean ± standard deviation. Discrete variables were reported as their frequencies and percentage of occurrences.

### Operative Procedure

The incision sites for the proposed MIS technique, as illustrated in Figure 1 (a), were summarized as follows:

**FIGURE 1.**
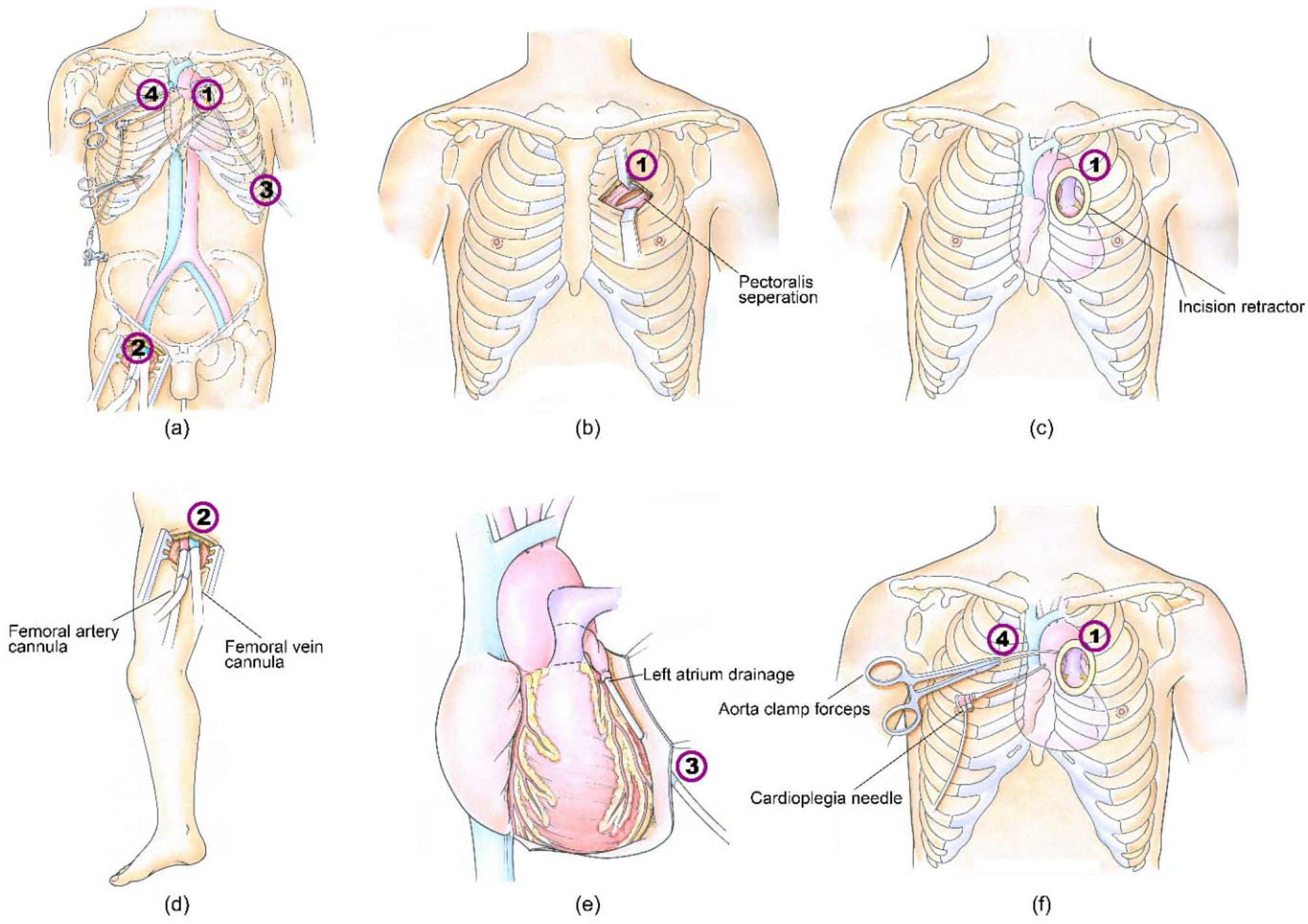
Artist depiction of (a) operative incisions in overall view for the proposed minimally invasive surgery, (b) incision ➀D, (c) the oval retractor for incision ➀D, (d) femoral cannulation via incision ➁, (e) left atrial appendage cannulation via incision ➂, and (f) clamping aorta and injecting cardioplegia via incision ➃.

- <u>Incision ➀:</u> the main incision for the surgical field; the upper border of the 3rd intercostal space between the left edge of the sternum and the midclavicular line, 3 cm in size for most patients (3-5 cm for older patients, i.e., 3 patients in this cohort were older than 10 years).
- <u>Incision ➁:</u> ∼1 cm above the inguinal ligament, 3 cm in size, used for femoral artery and vein access.
- <u>Incision ➂:</u> left 7^th^ intercostal space in the midaxillary line, 1 cm in size, used for left atrial appendage (LAA) drainage tubing and postoperative chest tubing.
- <u>Incision ➃:</u> right 2nd intercostal space close to the sternum edge, 1 cm in size, used for aortic occlusion clamp or superior vena cava cannulation.

The following section elaborates on the rationales and functions of these incisions.

### Cardiopulmonary Bypass Protocol

Incision ➀ is the main surgical field, which is a horizontal incision made by dissecting the pectoralis major muscle, along with its muscle bundle, and by incising the intercostal muscle as depicted in Figure 1 (b). During this step, special care is taken to prevent injury to the rib periosteum and mammary arteries. An oval retractor is then placed, as shown in Figure 1 (c).

The establishment of CPB requires incisions ➀, ➁, ➂, and ➃. In general, for patients older than 1 year of age, CPB is usually established by the femoral artery porous cannula (BIO-MEDICUS, Medtronic, Minneapolis, MN, USA) and a bipolar femoral vein cannula via the incision ➁, as shown in Figure 1 (d). Transesophageal echocardiography is used to facilitate placement of the bipolar vein cannula at the confluence of the persistent left superior vena cava and innominate vein. For the remaining patients (less than 1 year old), angiography is done preoperatively to determine the sizes of the femoral artery and vein, thereby confirming femoral catheterization feasibility. Access for patients with small or tortuous vessels is performed via incision ➃. The LAA tube is inserted into the LAA via incision ➂ and fixed with a 7-0 silk suture, as shown in Figure 1(e).

Then, CPB perfusion is initiated with a flow rate of 80-100 ml/kg-min at a pressure of 60-80 mmHg to achieve mild hypothermia at a nasopharyngeal temperature of 28-30°C. Vacuum-assisted venous drainage was applied with a negative pressure from -30 to -10 mmHg to assist in drawing blood from the femoral veins. The ascending aorta is clamped via incision ➃, right after the introduction3;, as shown in Figure 1 (f).

After total collapse of the patient’s heart, both the inferior vena cava (IVC) and SVC are snugged by using a Lark’s head knot or a surgeon’s knot with 7-0 silk suture, as shown in Figure 2. Particular attention is paid to ligating the IVC, because accessing the IVC during this step is challenging. It is imperative that the peripheral tissues surrounding the patient’s IVC are completely immobilized to ensure better access. Finally, innovative forceps are used to complete the ligation, as illustrated in Figure 2 (b).

**FIGURE 2.**
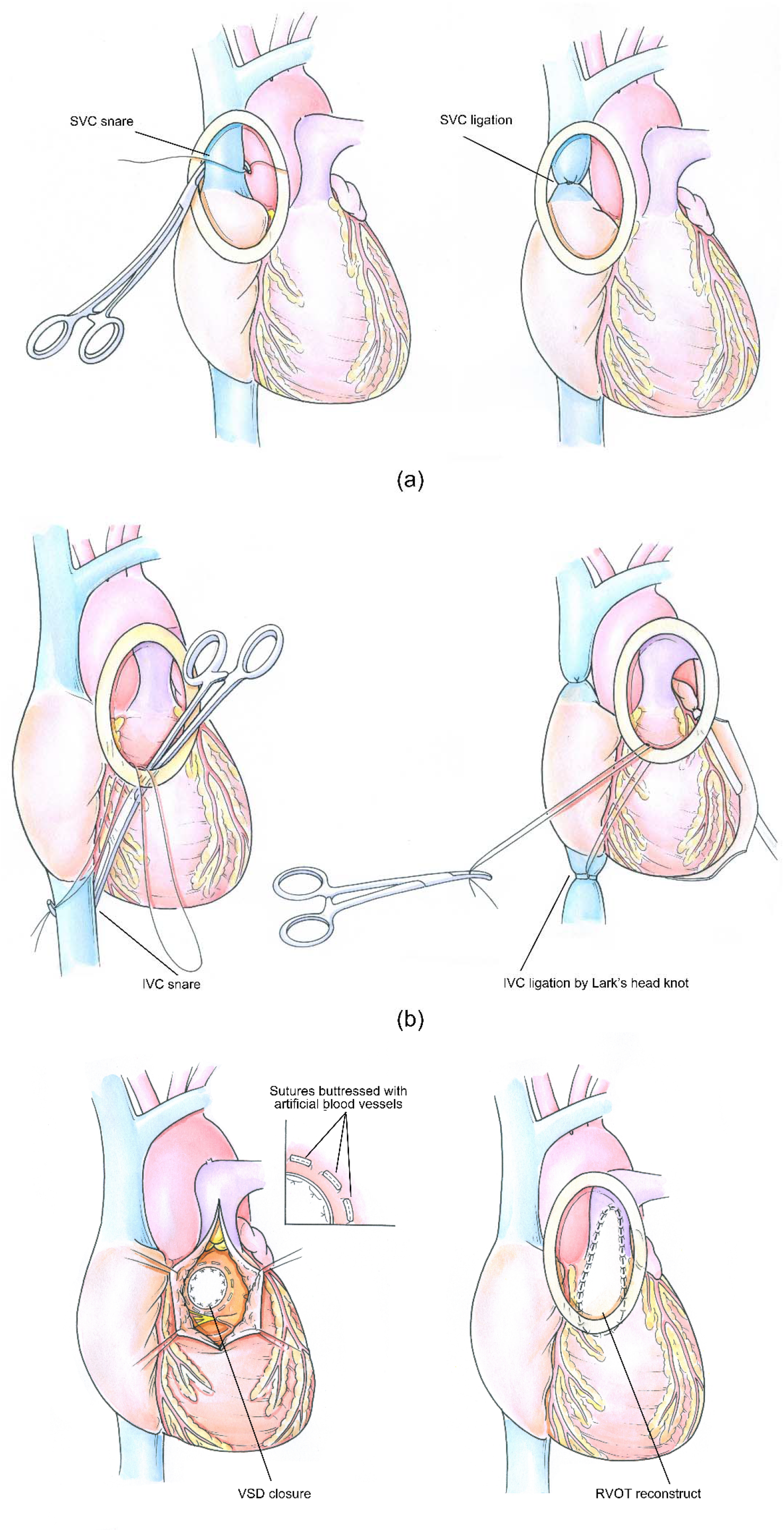
Artist depiction of (a) the ligation of the superior vena cava (SVC) by placing a vessel snare, (b) the ligation of the inferior vena cava (IVC) by placing a vessel snare, and (c) the procedure of repairing the ventricular septal defect (VSD) and reconstructing the right ventricular outflow tract (RVOT).

It should also be noted that prior to IVC and SVC ligation, the patient’s heart is perfused with autologous blood (10-15 ml/kg), with a potassium concentration of 26 mmol/L, a temperature of 30-32°C, and a hematocrit of 15%. Following ligation, the heart is recurrently perfused with oxygenated blood (10 ml/kg) with a potassium concentration of 9 mmol/L, a temperature of 28°C, and a hematocrit of 20%, at an interval of 15 to 20 minutes. Alternatively, it is also common practice to continuously perfuse the heart with histidine-tryptophan-ketoglutarate solution throughout the procedure. This commercial solution allows the surgical field to be ensanguined, but it may be a challenge to withdraw the solution from the femoral vein due to its low density.

### Intracardiac Procedure

The intracardiac procedure is similar to those for conventional heart operations for ToF. This section summarizes the key steps in our procedure. After the thymus is removed, the pericardium is opened 2 cm superior to the phrenic nerve. Great care should be taken to avoid harming the phrenic nerve in this step. Although the majority of the pericardium is suspended, a small portion might be resected and fixed with glutaraldehyde for later RVOT reconstruction, if necessary. Tissues that are disposed to RVOT stenosis, including hypertrophic muscle tissues on the crista supraventricularis, septal and parietal band, or any other ventricular wall, should be resected. The right ventricle is opened via a longitudinal incision. The ventricular septal defect closure is performed using a 0.4-mm-thick Gore-Tex patch (W. L. Gore Inc., Newark, DE, USA) and interrupted 4-0 or 5-0 mattress sutures buttressed with artificial blood vessels (1-2 mm). The RVOT was reconstructed with the autologous pericardium, as depicted in Figure 2 (c). If present, the patent foramen ovale or atrial septal defect should be closed via either the tricuspid valve or the atrium, depending on the surgeon’s preference. Once the repair is completed, the heart is de-aired, aided by negative pressure via the cardioplegia perfusion needle that remains at the aortic root. The aortic cross-clamp is then removed and is followed by sinus rhythm recovery. When the heart regains good contractility, the patient is weaned off bypass. At the conclusion of the procedure, incision ➀ was closed by intradermal running sutures after the intercostal nerve is blocked with lidocaine. The chest drainage tube is inserted through incision ➂. Specialized instruments designed to facilitate the completion of the proposed MIS procedure are shown in Figure 3. These instruments are now commercially available.

**FIGURE 3.**
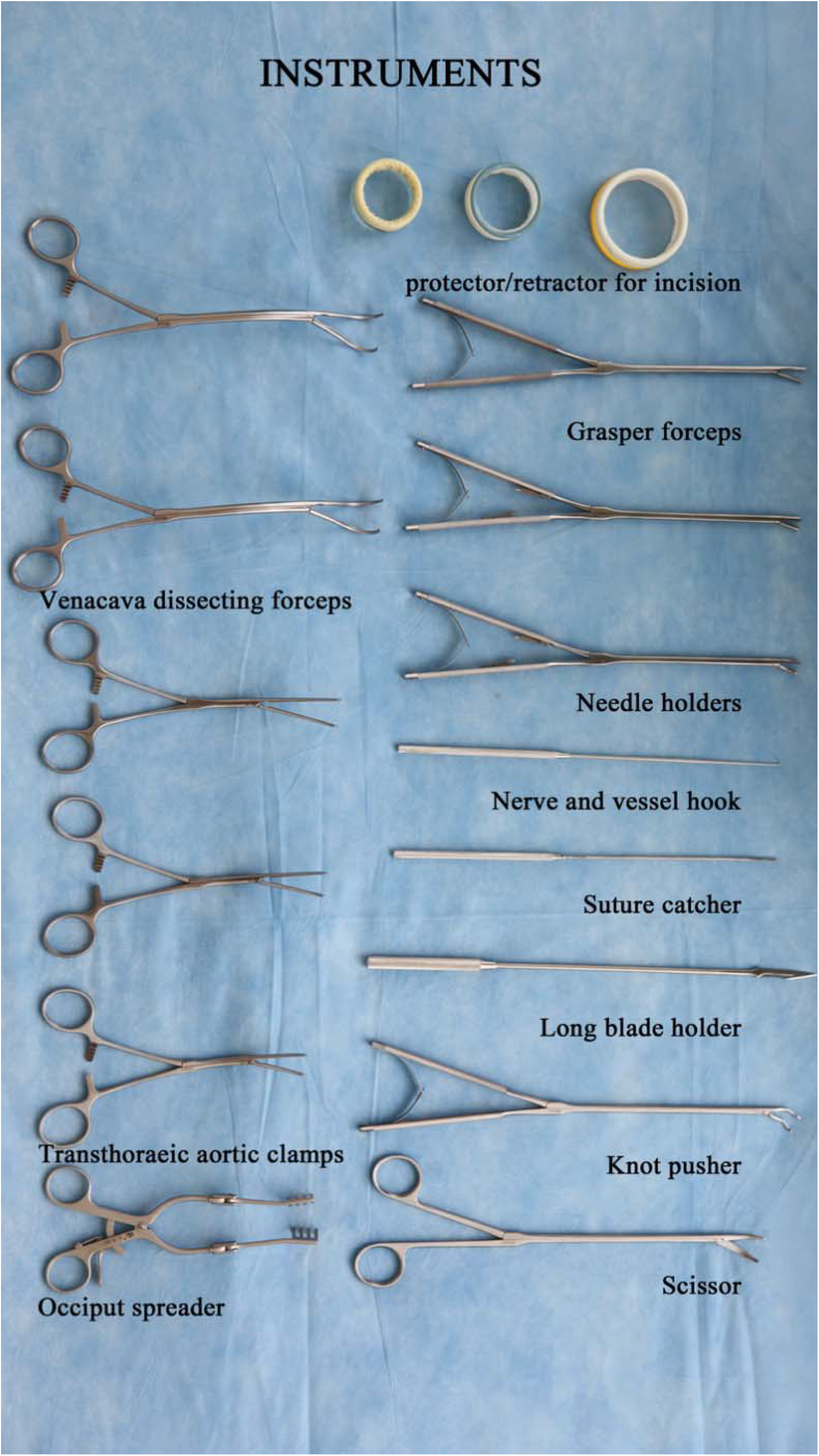
Specialized instruments developed to perform the proposed minimally invasive surgical procedure.

## RESULTS

### Patient Cohort

Table 1 summarizes baseline demographic and clinical data for all patients. Though most patients at baseline were classified as New York Heart Association functional class II, all patients were discharged as New York Heart Association functional class I. No events were found for bleeding, reoperation, conversion to open sternotomy, or any lesions to the femoral access site. Two cases utilized alternative cannulation instead of the femoral approach because (1) an infant patient had a small femoral artery with stenosis due to vasospasm, and (2) another infant patient had IVC malformation and a slender femoral vein within a collateral network.

**TABLE 1.** Demographic and baseline clinical characteristics.

| **Demographics** |  | **n = 105** |
| --- | --- | --- |
| Age (y) |  | 3±3.1 |
| ≤6 mo |  | 2 |
| 6 mo–1 y |  | 11 |
| >1 y |  | 92 |
| Males, n |  | 61 |
| Weight, kg |  | 12.8±6.3 |
| Height, cm |  | 95.1±23.4 |
| BSA, m<sup>2</sup> |  | 0.6±0.2 |
| Cardiovascular associations, n |  |  |
| Midcristal, infracristal, mVSD |  | 2, 16, 2 |
| PFO, PDA, pLSVC |  | 24, 11, 4 |
| Echocardiographic data |  |  |
| LV diameter, mm |  | 24.8±6.6 |
| LPA diameter, mm |  | 7±2.3 |
| RPA diameter, mm |  | 7±2.3 |
| McGoon |  | 1.8±0.5 |
| Nakata, mm<sup>2</sup>/m<sup>2</sup> |  | 158.8±81.7 |
| NYHA class |  |  |
| I/II |  | 16/89 |

All values are presented as median (range) or n (%).
*BSA*, body surface area; *LPA*, left pulmonary artery; *LV*, left ventricular; *mVSD*, muscular ventricular septal defect; *NYHA*, New York Heart Association; *PDA*, patent ductus arteriosus; *PFO*, patent foramen ovale; *pLSVC*, persistent left superior vena cava; *RPA*, right pulmonary artery.

Table 2 compared the patients’ laboratory data before and after the MIS procedure. The postoperative data indicated that all patients achieved favorable surgical outcomes. The procedural data were also tabulated in Table 2. Postoperative complications were present in 14 patients (13.3%):

**TABLE 2.** Pre- and postoperative laboratory examinations and interoperative surgical details.

| **Laboratory examinations (n = 105)** |  |  |  |
| --- | --- | --- | --- |
|  | Preoperation | Day of operation | Before discharge |
| COP, mmHg | 14.9±2.7 | 13.4±2.2 | 17.4±3.2 |
| SaO<sub>2</sub>, % | 88.4±8.7 | 81.7±3.5 | 100±0.3 |
| Systolic BP, mmHg | 94.2±7.5 | 92.3±8.9 | 96.1±6.1 |
| cLac, mmol/L | 1.1±0.6 | 2.3±0.9 | 1.7±0.9 |
| HgB, g/L | 135.9±26.6 | N/A | 113.9±16.2 |
| Hct, % | 40.1±7.2 | 29.6±6.2 | 34.2±4.7 |
| **Intraoperative details** |  |  |  |
| Transannular patch | 76 (72.4%) | Femoral artery/vein<br>cannula size, F | 10.4±2.3/ 14.7±1.9 |
| Aortic cross-clamp time,<br>min | 71.1±19.4 | Vecuronium/sufentanil, mg | 10±5.8/9.9±23.9 |
| Operation time, min | 223.1±42.1 | Hct in CPB, % | 29.6±6.2 |
| CPB time, min | 107.9±26.3 | Blood loss, ml | 15.7±5 |
| CPB temperature, °C | 27.7±4.1 | rScO<sub>2</sub>, % | 66.5±4 |
| *BP*, blood pressure; *cLac*, lactate concentration; *COP*, colloid osmotic pressure; *CPB*,<br>cardiopulmonary bypass; *Hct*, hematocrit; *HgB*, hemoglobin; *N/A*: not available; *rScO<sub>2</sub>*, regional<br>cerebral oxygen saturation; *SaO<sub>2</sub>*, oxygen saturation. |  |  |  |

- Two had left phrenic nerve palsy, leading to diaphragmatic elevation.
- Three had low cardiac output syndrome.
- One suffered a cerebral hernia with subarachnoid hemorrhage and died of acute stroke.
- One developed paroxysmal supraventricular tachycardia.
- Two developed subcutaneous emphysema.
- Five had pleural effusions.

A female, toddler patient with ToF combined with persistent left SVC died. The McGoon (1.5) and Nakata (94.6 mm^2^/m^2^) indices of this patient were at the lower end of the indices of our patient cohort. Pre-, inter-, and postoperative data showed that the surgery succeeded, although the CPB time was 128 min, which was relatively high in our cohort. The patient was in the ICU for 2 days and developed a subarachnoid hemorrhage. The patient’s symptoms were treated in a timely manner until the patient died of an acute stroke. Two patients with diaphragmatic elevation were left untreated. However, the elevation diminished (returned to a normal level) after 3 and 6 months. Otherwise, all patients recovered from complications under symptomatic treatment prior to discharge, with the exception of 1 patient who died. For example, the 3 patients with low cardiac output were treated by elevating the cardiac preload and administering a positive inotropic drug. Table 3 tabulates clinical details about the patients’ time in the ICU.

**TABLE 3.** Data collected during the stays in the intensive care unit.

| ICU details | Value (n=105) |  |  |
| --- | --- | --- | --- |
|  | Operative day | 1st Postoperative day | 2nd Postoperative day |
| Drainage, ml | 112.3±47.6 | 54.5±40 | 29.9±16.6 |
| HR, min<sup>-1</sup> | 133±16.2 | 131.5±15.9 | 135.3±14.5 |
| BP, mmHg | 92.3±8.9 | 88.4±8.5 | 87.7±8.4 |
| CVP, mmHg | 5.9±2 | 6.3±2.3 | 6.1±2 |
| DOP, µg/kg.min | 3.8±1 | ISO, ug/kg-min | 0.03±0.01 |
| DOP time, h | 122.9±86.3 | ISO time, h | 70.7±76.5 |
| SNP, µg/kg.min | 0.6±0.2 | Thoracic drainage time, d | 2.8±1.5 |
| SNP time, h | 46.8±54.9 | Ventilation time, h | 36.6±61.8 |
| EPI, µg/kg.min | 0.05±0.02 | ICU stay, d | 5.3±4.5 |
| EPI Time, h | 91.3±74.6 | Hospital stay, d | 22.1±9.9 |
| MIL, ug/kg.min | 0.5 (0.3-0.6) | MIL time, h | 105.2 (19-319) |

*BP*: blood pressure; *CVP*: central venous pressure, *DOP*: dopamine; *EPI*: epinephrine; *HR*: heart rate;431 *ICU*: intensive care unit; *ISO*: isoprenaline; *MIL*: milrinone; *SNP*: sodium nitroprusside.

Table 4 compares the key interoperative and postoperative clinical data from the current study with data from previously published studies for MIS of ToF. The comparisons show that the MIS technique used in the present study achieves acceptable outcomes.

**TABLE 4.** Comparison between the current study and previous studies for minimally invasive surgery for treatment of tetralogy of Fallot.

|  | Current Study | Lee et al. 2018 | Doshi et al.<br>2017 | Nicholson et<br>al. 2017 | Garg et al.<br>2001 |
| --- | --- | --- | --- | --- | --- |
| Main operative location | 3rd intercostal space | 4<sup>th</sup> intercostal space | 3<sup>rd</sup> intercostal space | Xiphoid | 3<sup>rd</sup> or 4<sup>th</sup> intercostal space |
| Main incision size | 3 mm for age ≤ 3; 5 mm for age ≥ 10; or 3-5 mm for the rest | 3.5-7 cm | 3-5 cm | 3.5-5 cm | NP |
| Cannulation technique | Femoral artery and vein | Aorta and systemic veins | Femoral artery and vein | Aorta and systemic veins | CCA, IVC, IJV |
| Sternal sparing | Yes | Yes | Yes | Sternal division | Transverse split sternotomy |
| Number of patients | All (n= 105) | 7 | 27 | 27 | 11 |
| Age, mo | 36±37.2 | 3.6 (1.2-4.8) | 13.2±9.77 | NP | 29.27 ± 9.18 |
| Weight, kg | 12.8±6.3 | 6(5-7) | 26.7±18.6 | NP | 12.1 ± 1.99 |
| Block time, min | 71.1±19.4 | NP | 48.33±15.73 | 44±16 | 76.27 ± 12.66 |
| Operation time, min | 223.1±42.1 | 292(226-324) | NP | NP | NP |
| CPB time, min | 107.9±26.3 | 144(93-163) | 83.66±26.65 | 85±33 | 113.54 ±<br>23.62 |
| Hospital stay,<br>d | 12.3±6.8 | 6(4-11) | 7±0.86 | 5.5±1.8 | 3.90 ± 0.53 |
| ICU stay, d | 5.3±4.5 | NP | 41.66±14.37<sup>‡</sup> | NP | 1.18 ± 0.40 |
| Drainage, ml | 213.9±144 | NP | 161.1±89.36 | NP | NP |

<sup>‡</sup> This number is copied directly from the corresponding citation. There may be a typo in this citation.
*CCA*, common carotid artery; *CPB*, cardiopulmonary bypass; *ICU*, intensive care unit; *IJV*, internal jugular vein; *IVC*, inferior vena cava; *NP*, not reported.

## DISCUSSION

Good exposure to the cardiac chamber is vitally important to the success of MIS. Unfortunately, conventional cannulation techniques often result in multiple tubes in the main surgical field. Therefore, a large incision was used in previous studies even for very young patients, i.e., 3-5 mm for patients less than 2 years of age.^16, 17^ This study proposed an innovative cannulation technique to obtain a tube-free main surgical field, leading to a relatively smaller incision size (3 mm for patients less than 3 years old). The key was to use the femoral cannulation technique, which keeps the left heart drain, the aortic clamp, and cardioplegia perfusion out of the main surgical field. The oval protective retractor further enhances clearance of the surgical field. It is worth noting that, because of the small sizes of the femoral artery and vein, negative pressure should be applied to achieve good hemodynamics during CPB.

The resulting tube-free main surgical field facilitates the completion of the MIS procedure without of the need for thoracoscopy. Also, the position of the main incision and the innovative CPB technique enable excellent access to the pulmonary arteries. The left atrium could be easily pressed down by pulling the left atrial drainage tube via incision l, leading to sufficient exposure to both the main and distal PA branches. Nevertheless, even with the tube-free surgical field, it can still be a challenge to ligate the IVC because it is located a significant distance from the surgical field. One of the most crucial steps is to ensure that the heart has sufficiently collapsed by applying suction pressure, lowering perfusion flow, and elevating the operating table for a short period of time. Subsequently, isolating the pericardial fold from the surrounding tissues could help the surgeon access the IVC ligation site. The special clamp with a flexible tip, as illustrated in Figure3, can also facilitate the ligation process.

Overall, the proposed MIS technique resulted in favorable surgical outcomes. Table 4 shows that the inter-and postoperative data obtained in the current study are within the acceptable ranges reported in the literature. The study at hand reported slightly longer lengths of stay in the hospital and in the ICU. However, this additional time was to be expected, because additional care and supervision were necessary for patients undergoing the novel surgical technique. Patients with ToF undergoing more recent MIS procedures in our center have had shorter stays, i.e., hospital stay = 6 days and ICU stay = 1 day, which are similar to 7.9±2.7 days of hospital stays for our open-heart operations. Innovative procedural considerations facilitated the quick recovery of the patients in our cohort. For instance, the main surgical incision was created along the pectoralis fascia to avoid primary damage to nearby muscles and ribs. The pectoralis major muscle was mobilized along the length of its muscle bundle. These special considerations could facilitate postoperative recovery, which is exemplified by no chest deformities in our cohort. Moreover, to avoid postoperative stenosis in the femoral artery and vein, we adopted a longitudinal incision and used a coronary bypass anastomosis to close the incisions—the first stitch 0.5 mm away from the edge of the incision and a 1-mm pitch of the needle. A lateral interrupted 1-0 silk suture was used to protect vessels in patients weighing less than 15 kg, whereas a longitudinal continuous 8-0 suture was routinely used for the remaining patients.

Offering generally favorable outcomes, the proposed MIS could be a safe option for early primary repair for patients with ToF. The benefit of early ToF repair by open-heart surgery is controversial. On the one hand, an early repair could be beneficial because it could reduce the pressure overload in the RV and the subsequent RV hypertrophy^22, 23^ and minimize secondary damage to other organ systems.^24, 25^ On the other hand, early repair with open-heart surgery is not widely used. The primary reason is the association between neonate repair and increased mortality and other burdens in perioperative care, including increased hospital stay, delayed sternal closure, and a high percentage of reoperations.^26-28^ Hence, MIS could be a good choice for early repair of ToF, and this study adds new evidence to support this idea. We performed early repair of ToF with the proposed MIS procedure in 2 patients (1 month and 5 months of age). It is worth noting that vessel cannulation was particularly challenging for these 2 patients because of the small size of their femoral vessels. Therefore, we have yet to apply this technique extensively for early repair of ToF. However, the alternative approach (described in the Methods section), whereby we intubated systemic veins via incision 4, could promote the early MIS repair of patients with ToF. Nevertheless, the successful completion of the 2 early repairs for our patients demonstrates the potential of the proposed MIS procedure to treat patients with a wide range of ages.

The authors acknowledge the limitations of this MIS technique. For example, it is known that a thoracotomy could damage the mammary gland.^13, 29^ We made special efforts to protect the mammary gland during our MIS procedure, especially for the female patients. Because most patients with ToF are infants or young children, the position of the mammary gland is largely unknown. In infants, the areola and breast mass lie on the 4th interspace in the infant and breast tissue, peripheral to the areolar border by as much as 1.5 cm.^30, 31^ Our minimal incisions avoid the future breast line and do not touch the 4^th^ rib, which is sufficiently remote from the areolar border. Nevertheless, predicting the location of the mammary gland in prepubertal^32^ patients is still a notable challenge. As such, further follow-up of breast development is necessary to ensure acceptable cosmetic and functional results. Additionally, though special instruments were developed for IVC ligation, accessing the IVC remains a challenge, because its ligation site is remote relative to the main surgical field. Special care should be taken to prevent injury to the IVC in the IVC ligating process, especially when the right ventricle is enlarged.

In conclusion, this study presents a single-center MIS experience with the largest cohort to date of patients with ToF. The proposed MIS technique allows a tube-free surgical field, and the overall outcomes are favorable. Our success in repairing 2 patients with ToF under 6 months of age is promising, indicating that this proposed technique may also be a good choice for earlier primary repair of patients with ToF.

## Data Availability

All data produced in the present study are available upon reasonable request to the authors

## ABBREVIATIONS

CHD: congenital heart diseases
CPB: cardiopulmonary bypass
ICU: intensive care unit
LAA: left atrial appendage
MIS: minimally invasive surgery
PFO: patent foramen ovale
RV: right ventricular
RVOT: right ventricular outflow tract
ToF: Tetralogy of Fallot

